# RAPID SEROLOGICAL TESTS HAVE A ROLE IN ASYMPTOMATIC HEALTH WORKERS COVID-19 SCREENING

**DOI:** 10.1101/2020.04.15.20057786

**Authors:** Angelo Virgilio Paradiso, SimonaDe Summa, Nicola Silvestris, Stefania Tommasi, Antonio Tufaro, Giuseppe De Palma, Angela Maria Vittoria Larocca, Maria Chironna, Vincenzo D’Addabbo, Donata Raffaele, Vito Cafagna, Vito Garrisi

## Abstract

Health workers are at high risk for SARS-CoV-2 infection and, if asymptomatic, for transmitting the virus on to fragile cancer patients. We screened 525 health workers of our Cancer Institute with rapid serological test Viva-Diag analyzingCOVID-19 associated-IgG/IgM. Six subjects (1,1%) resulted with Viva-Diag test not-negative for IgM. All 6 cases had RT-PCR SARS-CoV-2 test negative; repeating analysis ofIgG/IgM expression by CLIA assay also, 2 cases resulted IgM positive and 1 case IgG/IgM positive. This latter subject reported a contact with an infected SARS-CoV-2 person, a month earlier.In conclusion our study seems to suggest: a) a different analytical sensitivity inIgG/IgM evaluation for Viva-Diag and CLIA assays needing to be further determined; b) the ability of Viva-Diagrapid COVID-19 test to evidence health workers positive for Immunoglobulins expression. Discordant results of rapid serological tests with respect to RT-PCR stress the different clinical meaning the two assays can have, question clearly referring to further studies to optimize the utilization of rapid serological test in asymptomatic subjects at high risk for infection.

## Background

Recently, a novel coronavirus was first reported to be responsible for inter-person transmission of lethal pneumonia in China[1]. Subsequently, the cause of that transmissible disease was identified to be a new strain of the coronavirus family named SARS-CoV-2 and the associated disease was called coronavirus disease 2019(COVID-19) (2). COVID-19 rapidly spreadall over the world obliging governments to take extraordinary initiatives.

The actions rolled out to try to control the pandemic included establishing strict criteria to define patients from whom oropharyngeal swabs should be collected for molecular diagnosis of COVID-19(3). The RT-PCR test for the identification of viral nucleic acid is the current standard method for diagnosingthis disease.

However, despite these actions, worldwide cases of COVID-19 climbed above 1 million and deaths over 50,000 (4). From an analysis of this situation, experts have highlightedseveral critical issues.

Onefirst consideration concerns the role that asymptomatic infected persons, not routinely submitted to SARS-CoV2 RT-PCRtesting, could have in transmittingthe disease. Day (5)analyzed the figures from China and suggested that fourfifths of infected cases could be asymptomatic and Rothe confirmed that these cases canbe a source of infection transmission(6). Another point–which is directly related–regards the need to identifyhigh risk categories that may be exposed to infection so that they may be protectedand those that may beasymptomatic potential carriers of virus causingunexpected transmission. Apart from elderly individualsand those with co-morbidities (2), the category at greater riskof COVID-19 infectionis obviously represented byhealthcare workers(6).Anelli recently reported that in Italy, 20% of health workers could be infected(7).

Moving from the need to identifyunknown sources of infection and to adopt better rules for the diagnosis of COVID-19 in persons at high risk ofinfection, attempts have already been made to identify infected people in nursing facilities (8) and in hospitals (9) by using RT-PCRto test for SARS-CoV-2 positivity.

The RT-PCR test for the identification of viral nucleic acid is the current standard method for diagnosing COVID-19. However, this assay has some practical limitations (3) such as the annoying method to obtain biological material from the nasopharynx, the relatively long time to generate results andthe need for certified laboratories and specific expertise.

The Saw Swee Hock School of Public Health of the National University of Singapore recently reviewed(10) the diagnostic tests for COVID-19 infection currently undergoing clinical validation and also listed immunoassays detecting COVID-19 related IgG andIgM antibodies. This latter experimental attempt, based on previous experiences with epidemic viral SARS infection, argued that specific IgM antibodies against SARS-CoV-2 could be detected in blood after 3-6 days while IgGdetection occurred some days later (10). This approach is described as easy to handle and capable of providing results in 15 minutes (10).

Based on theinformation already reported,we considered the need to screen our healthcare personnel, whoare constantly on the front line working with immuno-depressed cancer patients,by utilizing an assay that could be applied in a large cohort of asymptomaticsubjects. To this purpose, we planned to screen all the health workers in our Cancer Institute for COVID-19 associated immunoglobulins with the rapid serological test,Viva-Diag^™^. Furthermore, to repeat testing all subjects with Viva-Diag^™^assaypositive forIgG and/or IgM, with standard RT-PCR and other serological tests was also planned.

The aims of the study were a) to verify the ability of the Viva-Diag^™^kit to identify subjects positive for COVID-19 relatedimmunoglobulins in an asymptomatic cohort of subjects, and b) to gather information on the percentage of healthworkers at true risk of virus exposure in a non-COVID-19 hospital.

## Material and methods

From March 26 to April 2, all the health workers employed at IstitutoTumori G. Paolo II,IRCCS, of Bari were invited to participate in a prospective trial in which venous blood samples were to be taken from allthe participating asymptomatic health workersand then submitted toarapid serological test,VivaDiag^™^. The study was approved by the Ethical Committee of the IstitutoTumori G. Paolo II, IRCCS, Bari with Protocol number CE 872/2020 together with the informed consent form which explained that quantitative serum immunoglobulin tests would be performed on the same blood samples and standard RT-PCR assays carried out on swab samples in caseswhere suspected infection was flagged up by the VivaDiag^™^ test.

A total of 525health workers replied and were enrolled.After signingtheir written informed consent form, all participantsfilled in a questionnaire which collected information on their possible risk ofCOVID-19 infection(contact with confirmed positive individualsor visits to areas with activeSARS-CoV-2 circulation). Venous blood samples were then collected andimmediately sent to the ClinicalPathologyLaboratory (Certified ISO-9001/2015; Head: E. Savino) and to the Institutional Biobank (Certified ISO-9001/2015; Head: A. Paradiso) of theIstitutoTumori G Paolo II, IRCCS, Bari (I) for Viva-Diag test performance.

The characteristics of the health workers who entered the study are reported in Table 1. Their median age was 48 years (range 20-73 yrs) and38%were male. A total of 56% of the enrolled workers were involved in direct clinical activities, 6% in laboratory practice, 8% in administrative activities and 30% in maintenance/cleaning activities. 1.4% of them reported minor clinical symptoms not directly referable to COVID-19 disease while 14% reported having had direct contact with individuals with suspected COVID-19 disease in the last two weeks.

**TABLE 1.** CHARACTERISTICS OF THE COHORT OF WORKERS SCREENED FOR COVID-19 DISEASE BY RAPID SEROLOGICAL TEST VIVA-DIAG^™^

| ***COHORT CHARACTERISTICS*** |  | ***N=525 (%)*** |
| --- | --- | --- |
| ***Sex*** |  |  |
|  | Male | 199 (37.9%) |
|  | Female | 326 (62.1%) |
| ***Age*** |  | 48years (range: 20-73) |
| ***Role*** |  |  |
|  | Clinical activity | 294 (56%) |
|  | Laboratory | 34 (6.5%) |
|  | Amministrative | 40 (7.6%) |
|  | Mainantenance/cleaning | 157 (29.9%) |
| ***Subjects with SARS-CoV-2 contacts*** |  | 71 (13.5%) |
| ***Subjects with minor symptoms*** |  | 7 (1.4%) |

### SARS-CoV-2 rapid IgG-IgM Test

SARS-CoV-2 rapid IgG-IgM combined antibody test kit, Viva-Diag^™^, was designed and manufactured by Jiangsu Medomics Medical Technologies, Nanjing-China. It is a lateral flow qualitative immunoassay for the rapid determination of the presence or absence of both anti-SARS-CoV-2-IgM and anti-SARS-CoV-2-IgG in human specimens (whole blood, serum, and plasma). A surface antigen from SARS-CoV-2 which can specifically bind to SARS-CoV-2 antibodies (including both IgM and IgG) is conjugated to colloidal gold nanoparticles and sprayed onto conjugate pads. The SARS-CoV-2 rapid IgG-IgM combined antibody test strip has two mouse anti-human monoclonal antibodies (anti-IgG and anti-IgM) stripped on two separated test lines. The presence of SARS-CoV-2 IgG and IgM antibodies is indicated by a red/purple line that appears in the specific region for those antibodieson the device. If the specimen did not contain SARS-CoV-2 antibodies, no labeled complexes were bound. Each test was evaluated by two operators and a picture was taken of the result. In case of disagreement between the two operators, the picture was evaluated by a third party.

### Molecular detection of SARS-CoV-2

In cases whereCOVID-19 was suspected based on the Viva-Diag^™^test results(n=6), oropharyngeal swabs were collectedon the following day for standard SARS-CoV-2 RT-PCR testing. The RT-PCR tests were immediately performed at the Laboratory of MolecularEpidemiology and Public Health(Head: M.Chironna) of the University of Bari (I). The swabs were subjected to nucleic acid extraction by MagNA Pure (Roche Diagnostics, Mannheim, Germany), according to the manufacturer’s instructions. The presence of the E gene, RdRP gene and N gene of the SARS-CoV-2 virus were identified by a commercial real-time PCR assay (Allplex 2019-nCoV Assay; Seegene, Seoul, Republic of Korea). Samples were considered positive at molecular screening if all the three genes were detected. The WHO Real-time-PCR protocol was used to confirm the presence of SARS-CoV2 (https://www.who.int/docs/default-source/coronaviruse/uscdcrt-pcr-panel-for-detection-instructions.pdf?sfvrsn=3aa07934_2).

### Chemiluminescence (CLIA) IgG/IgM detection

The IgG/IgM dosage was determined utilizing MAGLUMI^™^ 2019-nCoV IgG (Cat. Ref. 130219015M) andIgM (130219016M) (CLIA) according to the manufacturer’s indications (Shenzhen New Industries Biomedical Engineering Co., Ltd; www.snibe.com). Briefly, the kit is designed forindirect chemiluminescence qualitative-semiquantitative in vitro immunoassayof IgG and IgM antibodies against SARS-CoV-2in human plasma and serum.The pre-diluted biological sample, buffer and magnetic microbeads coated with recombinant SARS-CoV-2 antigen were mixed and incubated to create immunocomplexes.After magnetic field exposure and repeated washing,IgG and IgM antibodies labeled with ABEI were added;the chemiluminescence starter activated the lightreaction which was detectedby the photomultipier BIOLUMI^™^ 8000 (www.snibe.com). Results were expressed asrelative light units (RLU). The cut-off value of the test was determined as the mean luminescence value of normal sera plus 5 folds of SD. Results were considered positive if the signal/cutoff (S/C) ratio was ≥ 1.

### Statistical analyses

A descriptive analysis of the VivaDiag^™^test results with respect to the health workers’ characteristics is shownin Table 1.VivaDiag^™^ test resultsare compared to the other laboratory assay results in Table 2.

**Table 2.** CHARACTERISTICS OF SUBJETCS WITH NOT NEGATIVE COVID-19 VIVA-DIAG TEST RESULT AND COMPARISON WITH OTHER ASSAYS

| ID | Age<sup>1</sup> | Activity | 2019-nCoV contacts | Minor symptoms | ViVaDiag™ Test Result |  | SARS-CoV-2 RT-PCR | CLIA ANALYSIS |  |
| --- | --- | --- | --- | --- | --- | --- | --- | --- | --- |
|  |  |  |  |  | IgM<sup>2</sup> | IgG<sup>2</sup> |  | IgM (AU/mL) | IgG (AU/mL) |
| #1 | Older | Not-Clinical | No | No | Weak | Neg | Neg | 1.715* | 0.172 |
| #2 | Older | Clinical | Yes | No | Pos | Neg | Neg | 1.130* | 0.132 |
| #3 | Older | Clinical | Yes | No | Weak | Neg | Neg | 0.492 | 0.390 |
| #4 | Older | Not-Clinical | No | No | Weak | Neg | Neg | 0.569 | 0.150 |
| #5 | Younger | Not-Clinical | No | No | Weak | Neg | Neg | 0.826 | 0.283 |
| #6 | Older | Clinical | Yes | No | Pos | Pos | Neg | 1.184* | 6.918* |

\*cut-off for positivity at CLIA >1 AU/ml;<sup>1</sup>Older and younger indicate an age greater or lower, respectively, than median age of the cohort (48 years); <sup>2</sup>Weak indicate a low-intensity band; Pos: positive result; Neg: negative result.

## RESULTS

A total of 85% of all the health workers in our National Cancer Research Center, IstitutoTumoriG. Paolo II inBari, Italy were enrolled. Those who did not takepart inthe study were equally distributed among the various work categories. About 92% of the participants had routine daily contacts with clinical departments.

In 6 out of 525 cases, the VivaDiag^™^ test(Table 2) provided results that were not negative (weak or strong staining) in the IgM band; in one case, the test described a strong IgG/IgM intensity.Three cases, comprised the latter one, werehealth workerswho reported had had recent contacts with COVID-19 patients. None of them presented clinical symptoms associated with COVID-19 disease. Only one was younger than 55 years of age.

The day after their VivaDiag^™^ test, oropharyngeal swabs were collected from all 6 subjects for SARS-CoV-2 RT-PCR testing. None of their test results were positive for the virus (Table 2).

In order to gain further insights into theIgG/IgManalytical sensitivity provided by the colorimetric VivaDiag^™^kit, a plasma aliquot(from the same Viva-Diag test blood sample) of all 6 subjects was utilized for CLIA analysis of IgG/IgM. The CLIA results showedIgM positivity in two cases (a nurse and a member of the hospital’s cleaning staff, Table 2) and the confirmation of the strong positivity for IgG/IgM in the third one.

## DISCUSSION

Asymptomatic infection with SARS-CoV-2 has been highlighted to beone of the major sources for the spread ofCOVID-19 and may be the cause for the escalation of casesall over the world. Pioneering efforts in some countries to screen asymptomatic people showed that the problem is not easy to handle. In the small town of Vo’Euganeo(a red zone quarantined area for incident COVID-19 casesin northern Italy), all 3300 inhabitants, including asymptomatic individuals, were tested bystandard RT-PCR in the attempt to identifySARS-CoV-2 carriers(11). Six asymptomatic individualswere found to be positive (0.2%).In Iceland, a biotech company working on behalf of the country’sChief Epidemiologist screened the general population. Preliminary results reported that more than 99% of the inhabitantstested had negative results (12).These findingsare in contrast with recent figures from China supporting the hypothesis that four fifthsof SARS-CoV-2 infected subjects could be asymptomatic (5). In sum, while the trueprevalence of asymptomatic infected persons is a matter of debate, it has definitely been accepted that they represent one of the major contributors in thespread of the disease.

Although those studies did not provide information on specific risk categories, health workers category are obviously considered a category at higher risk ofinfection even when they are involved in general practices or in non-COVID-19 specific hospitals(7).As a consequence, the problem of detecting asymptomatic SARS-CoV-2 infected health workers needs to be considered one of primary relevance.Policies should be enacted to specificallyprotecthealthcare workers and stymie circulation of the virus inthe sensitivecontexts in which they work.

In order to gain insight into this aspect of the epidemic, we tested525 asymptomatic workers of our Cancer Institute where immuno-depressed patients are extremely frequent and sporadic COVID-19 positive cases had recently been reported. SARS-CoV-2-associatedimmunoglobulinswere assayed by utilizing a rapid serological colorimetric test.

We believe the results we obtained to beof great interest. The VivaDiag^™^test was able to find about 1,1% of cases with suspected presence of COVID-19 specificimmunoglobulins. This figure can be considered in line with the percentage of asymptomatic COVID-19 positive cases found in other population-wide experiences. The only two reports regarding healthcare personnelare fromThe Netherlands (9) andfrom a nursing-core facilities in Washington, U.S.A. (8). In the Netherlands, Reuskentested 1097 health workerswith mild respiratory complaints from nine hospitals and found that 4.1% of them were positive.Kimball analyzed residents of a longterm care skilled nursing facility and reported that 30% of them tested positive. Since those two studies involved subjects and facilities with very different characteristics with respect to those of our Cancer Institute, the present experience can be considered to be the first to address the problem of screening asymptomatic health workers for SARS-CoV-2 infection.

In our study, none of the six workers that tested positive for COVID-19 with the VivaDiag^™^kithad oropharyngeal swabspositive forSARS-CoV-2 upon RT-PCR testing. After one week of follow-up,all six operatorswere still asymptomatic. Our finding that subjects who were positive for COVID-specific IgM/IgG did not present any viral load is in agreement with Lou (13) who reported that higher Ig levels can be asynchronous with respect to viral presence and that can persist long time aftervirus RT-PCR testsbecome negative.One more point that drew our attention wasthe difference between VivaDiag^™^and CLIA in detecting the twoimmunoglobulins. SNIBE (www.SNIBE.com) recently announcedthat its MAGLUMI SARS-CoV-2 IgM/IgG Kits receivedits CE mark. Considering the cases that tested positivewith theVivaDiag^™^kitand negative withthe CLIAMAGLUMIkit, it would appearthatthe VivaDiag^™^testcould have a greater analytical sensitivity. However, our study was not designed to directly compare the performances of the two assays and, then, we cannot exclude conversely a better specificity of CLIAMAGLUMI kit. This because negative cases at VivaDiag^™^kit were not tested with the CLIAassay. This aspect will be further investigated in ournext trialin which all cases will be double,Viva-Diag and CLIA MAGLUMI kits, tested.

Separate discussion deserves the case strongly positive for IgG/IgM at both serological tests. The subject was a nurse who referred contacts with a COVID-19 patient 30-days before and then it is easy to relate herIg positivity to that event. It is the first time that a screening with rapid serological test individualize a single asymptomatic health worker with presumed passed virus infection within a cohort of hundreds of subjects.

## CONCLUSIONS

Our study seems to suggest that: a) rapid serological tests can have the ability to individualize subjects who had a previous contact with SARS-CoV-2 virus; b) apercentage, albeit very small,of our asymptomatic health workers could have been previously exposed tothe virus. Further studies are required to confirmand fully interpret our findings and to propose an optimal utilization of serological test in the clinics.

## Data Availability

All data are presented in the paper

## Notes

### Competing Interest Statement

The authors have declared no competing interest.

### Funding Statement

Italian Ministry of Health (Ricerca Corrente 2020)

